# Hepatitis E virus seroprevalence in South Africa from a multi-site study among HIV-negative and HIV-positive adults and age-stratified children (2–17 Years)

**DOI:** 10.64898/2026.05.01.26352167

**Authors:** Tarun Saluja, Nigus Telele, Elizabeth Hellström, Essack Mitha, Maphoshane Nchabeleng, Rita Baiden, Naveena Aloysia D’Cor, Sridhar Vemula, Ju Yeon Park, Lei Yang, Jiyoung Lee, Deok Ryun Kim, Sunju Park, Sanet Aspinall, HuiRong Pan, J Wai-Kuo Shih, Julia Lynch

**Author notes:** Corresponding author: Nigus Telele.

## Abstract

**Background:** Hepatitis E virus (HEV) seroprevalence varies by age and geography. Data on HEV seroprevalence across age groups and among people living with HIV (PLWH) in South Africa is scarce.

**Methods:** We conducted a prospective multi-site assessment of anti-HEV IgG seroprevalence on 859 South African participants enrolled at three clinical research centres including Newtown Clinical Research Centre in Johannesburg, Be Part Research in Mbekweni, Paarl, Western Cape, and Mecru Clinical Research Unit in Garankuwa, Pretoria. Participants comprised adults aged 18 – 45 years (PLWH, n = 178 and HIV-negative, n = 232), and children aged 2–17 years (n = 449). Anti-HEV IgG serostatus and antibody titer were measured using a commercial ELISA kit and a WHO reference standard. Seroprevalence was assessed by site, age group, sex, and HIV status.

**Results:** Overall anti-HEV IgG seroprevalence was 18.0% (95% CI: 15.6–20.8). Adults had the highest seroprevalence (27.3% among all adults; 29.2% among PLWH and 25.9% in HIV-negative adults), while adolescents aged 12–17 years had the lowest (6.9%), and young children aged 6–11 years and 2–5 years had 10.3% and 13.0%, respectively. Adults had significantly higher odds of seropositivity than children (aOR 2.8, 95% CI: 1.5–5.5, *p* = 0.002). A significant site-specific variation was also observed among healthy adults and adolescents: Newtown Clinical Research Centre (23.0% and 14.0%) and Be Part Research (34.5% and 7.3%) had higher seroprevalence compared with those from Mecru Clinical Research Unit (17.2% and 1.5%, *p = 0.0499* and *0.0262*, respectively). A higher mean antibody titer observed in younger children aged 2–5 years (5.06 IU/mL), compared with adults (0.88 IU/mL among PLWH and 0.68 IU/mL among HIV-negative adults), and with older children (2.02 IU/mL in those aged 6–11 years and 0.67 IU/mL in those aged 12–17 years).

**Conclusions:** HEV seroprevalence in South Africa was highly heterogeneous, varying markedly by age group and study site. These findings highlight the need for strengthened, integrated HEV surveillance to better define transmission patterns and to inform evidence-based considerations for prevention of infection.

## 1. Introduction

Hepatitis E virus (HEV), identified in the early 1980s, is one of the most common global causes of acute viral hepatitis (1). The highest global disease burden is observed in regions with limited access to clean drinking water, as fecal contamination of water sources remains a primary mode of transmission (2, 3). HEV consists of four primary genotypes (1–4) that infect humans. Genotypes 1 and 2 are exclusively human pathogens and are primarily spread through the fecal-oral route, often triggering large-scale outbreaks in areas with inadequate sanitation (4). On the other hand, genotypes 3 and 4 are zoonotic, linked to the consumption of undercooked meat or direct contact with infected animals, and are more commonly found in developed regions (5).

While HEV infection is often self-limiting in immunocompetent individuals, it can cause severe complications in vulnerable populations, including immunocompromised individuals and pregnant women (6, 7). In people living with HIV (PLWH) with low CD4+ T-cell counts and impaired immune responses, HEV can persist, leading to chronic hepatitis and progressive liver fibrosis (8). Studies indicated that HEV seroprevalence is higher among HIV-infected individuals compared to the general population (9, 10) and HEV-HIV co-infection may accelerate progression to liver cirrhosis, complicating clinical management (11).

In addition, children represent a vulnerable population affected by HEV though traditional serological tests may lack diagnostic precision, especially in areas with high endemicity (12, 13). Although HEV infection is usually self-limiting in immunocompetent children, those with underlying liver diseases, or immunocompromised children, are at higher risk for severe, prolonged or recurrent infections (13, 14). In the pediatric population, diagnosis can be difficult due to nonspecific symptoms, and the disease often remains underdiagnosed due to its overlap with other childhood illnesses (15). Serological tests detecting anti-HEV IgM and IgG antibodies are used for diagnosis, but routine screening is not typically performed (16). Importantly, HEV infection during pregnancy is also associated with substantially increased maternal morbidity and mortality (17, 18).

In South Africa, HEV is endemic, and a high seroprevalence has been reported (19-23). The studies suggest a mix of waterborne and zoonotic transmission, with increasing genetic diversity of the virus detected in pig products (23). However, routine HEV screening is not a standard practice in South African healthcare, which results in significant underreporting and a lack of comprehensive surveillance data (22). Early studies conducted in the 1990s reported HEV seroprevalence rates among the South African population ranging from 2% to 10% (21, 22), but these early estimates were limited by small sample sizes and regional focus.

Recent studies indicate a rising burden of HEV infection, with seroprevalence rates increasing, though they vary considerably across different provinces. In the Western Cape, a seroprevalence of 27.9% has been reported, with higher rates observed among individuals over 30 years of age, and with pork and bacon/ham consumption suggested as a risk (19), suggesting zoonotic transmission may be a factor.

Similarly, in the Free State, seroprevalence was significantly higher at 60.9%, highlighting substantial past exposure in certain populations (20). These findings suggest that HEV is more prevalent than previously recognized, but its public health importance remains underestimated. Despite the increasing evidence of widespread HEV exposure in South Africa, data specific to high-risk populations, such as immunocompromised individuals and children, are still limited as most available studies have been conducted in adult or mixed-age populations without detailed age-specific estimates, highlighting an important gap in understanding early-life transmission dynamics.

Therefore, the objective of this study was to determine the seroprevalence of anti-HEV IgG antibodies among individuals aged 2–45 years enrolled in a vaccine trial across three clinical research sites in South Africa. In addition, the study aimed to compare seroprevalence across key demographic subgroups, including geographic location, age, sex, and HIV status, and to quantify anti-HEV IgG titers among seropositive participants using a WHO-standardized reference.

## 2. Materials and Methods

### Study Population

Baseline blood samples were collected from participants enrolled from three clinical trial sites in South Africa participating in a Phase 2b, open-label study to evaluate the immunogenicity and safety of Hecolin® in HIV positive/negative adult participants and children (ClinicalTrials.gov ID: NCT06306196). They were stratified by age into four different cohorts: 18–45 years, 12–17 years, 6–11 years, and 2–5 years. The adult cohort was further stratified into HIV-positive and HIV-negative groups. The trial sites include Be Part Research (Mbekweni, Paarl in Western Cape), Newtown Clinical Research Centre (in Johannesburg), and Mecru Clinical Research Unit (in Garankuwa, Pretoria).

The Newtown Clinical Research Centre is a clinic-based research centre in urban Johannesburg. The Be Part Research site is located in Mbekweni, Drakenstein sub-district of peri-urban Cape Town. MeCRU (Mecru Clinical Research Unit) is a university-based clinical trials site at Sefako Makgatho Health Sciences University (SMU) based in Garankuwa in peri-urban Pretoria. All sites draw participants of lower socioeconomic status. Participants were recruited for the trial from their communities through outreach activities, referrals from local health services, and direct engagement with past volunteers.

### Sample Collection and Processing

Approximately 3.5 mL blood sample was collected from each participant prior to vaccination using a Gold-Top serum separator tube (SST). The blood samples were centrifuged within one hour of collection at 1400g for 10 minutes at the clinical sites and shipped refrigerated to Cytespace Africa Laboratories, a clinical laboratory in Pretoria, on the same day. Serum was separated, aliquoted, and stored at −70°C until analysis at Cytespace Laboratories. Before enrollment, adult participants were tested for HIV at the respective clinical sites. Participants with a prior HIV diagnosis were required to be on stable antiretroviral (ARV) therapy, and newly diagnosed participants were required to have received ARV for at least four weeks prior to enrolment.

### HEV Serological Testing

Anti-HEV IgG was measured using the WANTAI HEV IgG ELISA kit (Beijing Wantai Biological Pharmacy Enterprise, Beijing, China) following the manufacturer’s instructions. Anti-HEV IgG seropositive samples were further tested with a quantitative ELISA developed in-house using a WHO Reference Reagent for antibodies to hepatitis E virus, human serum (NIBSC code: 95/584) (24, 25).

Additionally, for HIV-positive participants, HIV viral load was measured using the GeneXpert platform with a detection limit of 40 copies/mL, while CD4% and absolute CD4 counts were determined using the BD FACSCanto II flow cytometer. All assays were conducted according to the manufacturer’s instructions and standard laboratory protocols to ensure precise and reliable results.

### Statistical analysis

All data were entered into an electronic data capture system, and analyses conducted using SAS 9.4 (SAS Institute, Cary, NC). Descriptive statistics were used to summarize demographic characteristics and anti-HEV IgG seroprevalence rates at baseline (before vaccination). Seroprevalence was compared across age groups, gender, geographic location of the clinical sites, and potential risk factors such as HIV status, viral load, and CD4 cell count. Categorical variables were analyzed using chi-square or Fisher’s exact tests, while continuous variables were assessed using t-tests or Mann-Whitney U tests for two group, ANOVA or Kruskal–Wallis test for more than two groups as appropriate. Logistic regression analysis was conducted to identify independent predictors of anti-HEV IgG seropositivity, adjusting for relevant covariates. All statistical tests were two-sided with a significance level of 5%.

Ethical approval was obtained from the relevant authorities including South African Health Products Regulatory Authority (SAHPRA Ref No 20231106 and 20250213), Pharma Ethics, South Africa (Ref No. 2311236044), Sefako Makgatho Health Sciences University Research Ethics Committee (Ref No SMUREC/M/523/2023 CR), and the Institutional Review Board of the International Vaccine Institute, South Korea (IRB NUMBER: 2023-010). Informed consent was obtained from all adult participants and the legal guardians of minor participants.

## 3. Results

A total of 955 participants were screened for eligibility in the trial, of whom 95 did not meet the inclusion criteria, and 860 participants were enrolled across three sites in South Africa (Figure 1). All participants provided baseline blood samples prior to vaccination, except one who withdrew before sample collection. This resulted in a total of 859 participants included in the analysis across the three sites: Be Part Research (Mbekweni in Western Cape) and Newtown Clinical Research Centre (in Johannesburg) each enrolled 343 participants, while Mecru Clinical Research Unit (Garankuwa in Pretoria) enrolled 173 participants.

**Figure 1.**
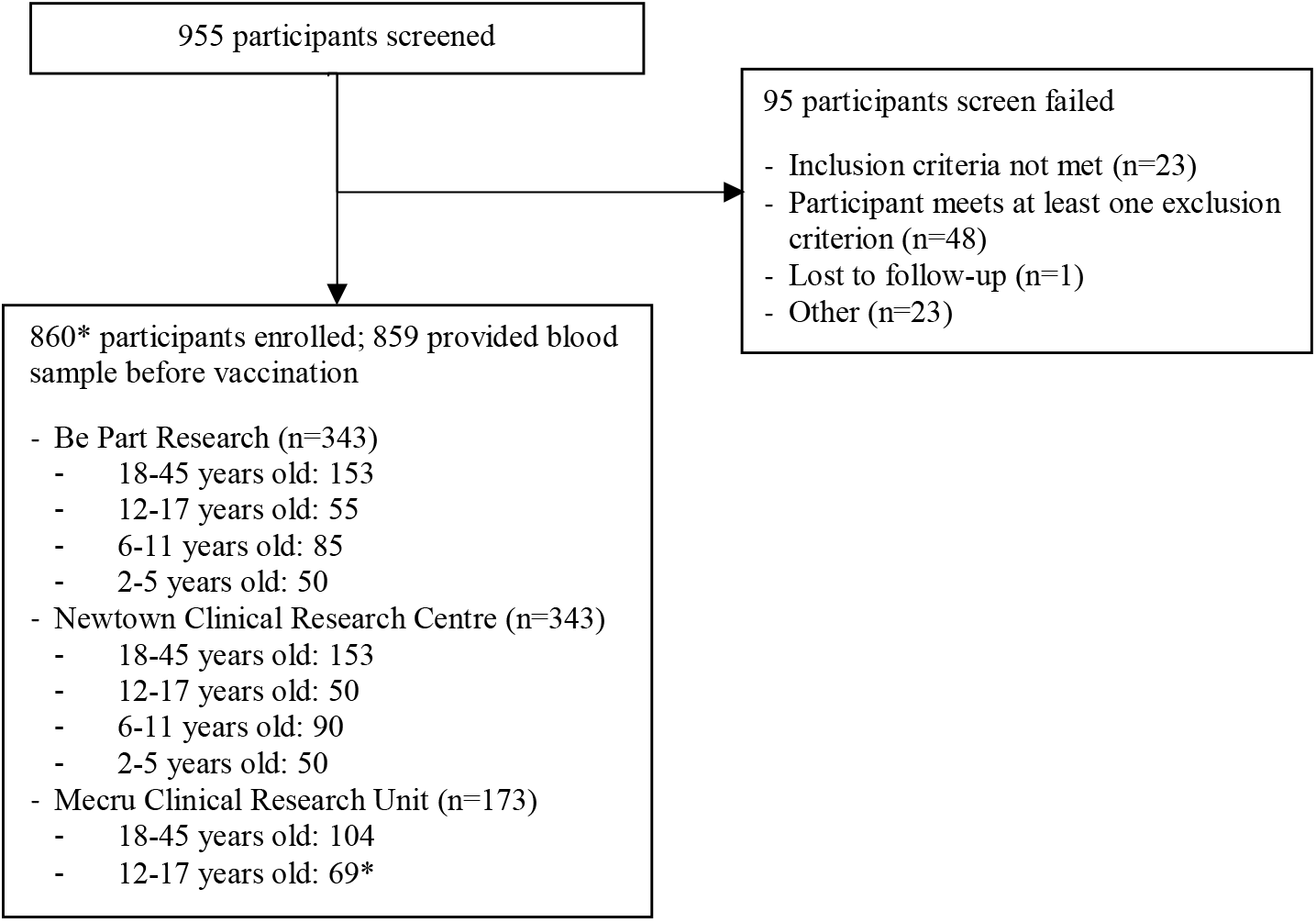
Flowchart illustrating participant disposition. ^*^ 1 participant from Mecru Clinical Research unit in the 12-17 years old age group withdrawn before blood drawn and is excluded from analysis

### 3.1. Demographic and Clinical Characteristics

The study population included adults living with HIV (PLWH, n = 178), HIV-negative adults (n = 232), and healthy participants from three pediatric cohorts (12–17 years, n = 174; 6–11 years, n = 175; 2–5 years, n = 100). Summaries of the demographic and clinical characteristics of participants stratified by HIV status and age group are presented below in Table 1.

**Table 1.** Demographic and clinical characteristics by age group and HIV status.

| Characteristics | Total<br>(n=859) | PLWH*<br>18-45 years<br>(n=178) | 18-45 years<br>(n=232) | Healthy participants<br>12-17 years<br>(n=174) | 6-11 years<br>(n=175) | 2-5 years<br>(n=100) |
| --- | --- | --- | --- | --- | --- | --- |
| **Site** | **n (%)** |  |  |  |  |  |
| Be Part Research | 343 (39.93%) | 66 (37.08%) | 87 (37.50%) | 55 (31.61%) | 85 (48.57%) | 50 (50.00%) |
| Newtown Clinical Research Centre | 343 (39.93%) | 66 (37.08%) | 87 (37.50%) | 50 (28.74%) | 90 (51.43%) | 50 (50.00%) |
| Mecru Clinical Research Unit | 173 (20.14%) | 46 (25.84%) | 58 (25.00%) | 69 (39.66%) | 0 (0.00%) | 0 (0.00%) |
| **Gender** | **n (%)** |  |  |  |  |  |
| Male | 401 (46.68%) | 58 (32.58%) | 98 (42.24%) | 97 (55.75%) | 96 (54.86%) | 52 (52.00%) |
| **Race** | **n (%)** |  |  |  |  |  |
| Black | 838 (97.56%) | 177 (99.44%) | 228 (98.28%) | 171 (98.28%) | 166 (94.86%) | 96 (96.00%) |
| Coloured | 20 (2.33%) | 1 (0.56%) | 4 (1.72%) | 3 (1.72%) | 9 (5.14%) | 3 (3.00%) |
| Indian/Asian | 1 (0.12%) | 0 (0.00%) | 0 (0.00%) | 0 (0.00%) | 0 (0.00%) | 1 (1.00%) |
| **Age (years)** |  |  |  |  |  |  |
| Mean (SD) | 19.52 (12.13) | 33.96 (6.76) | 27.62 (7.61) | 14.17 (1.64) | 8.50 (1.66) | 3.68 (1.00) |
| Median | 16.00 | 35.00 | 26.00 | 14.00 | 8.00 | 4.00 |
| Min, Max | 2.00, 45.00 | 19.00, 45.00 | 18.00, 44.00 | 12.00, 17.00 | 6.00, 11.00 | 2.00, 5.00 |
| **BMI Index (kg/m<sup>2</sup>)** |  |  |  |  |  |  |
| Mean (SD) | 21.48 (6.43) | 26.61 (6.05) | 25.27 (6.29) | 19.86 (3.52) | 16.13 (2.12) | 15.75 (1.27) |
| Median | 19.70 | 25.70 | 23.30 | 19.25 | 16.00 | 15.80 |
| Min, Max | 10.30, 39.80 | 15.90, 39.70 | 16.30, 39.80 | 12.80, 37.70 | 11.50, 24.40 | 10.30, 18.30 |
| **Viral Load†** |  |  |  |  |  |  |
| Undetectable (<40 copies/mL) | - | 162 (91.01%) | - | - | - | - |
| Detectable (≥40 copies/mL) | - | 16 (8.99%) | - | - | - | - |
| **Viral Load‡ (copies/mL)** |  |  |  |  |  |  |
| Mean (SD) | - | 2427.94 (5956.16) | - | - | - | - |
| Median | - | 201.00 | - | - | - | - |
| Min, Max | - | 55.00, 24000.0 | - | - | - | - |
| **CD4 Categories§** |  |  |  |  |  |  |
| SIM | - | 0 (0.00%) | - | - | - | - |
| MIS | - | 25 (14.04%) | - | - | - | - |
| NIS | - | 153 (85.96%) | - | - | - | - |
| **CD4 Count¶ (cells/uL)** |  |  |  |  |  |  |
| Mean (SD) | - | 843.40 (342.75) | - | - | - | - |
| Median | - | 792.50 | - | - | - | - |
| Min, Max | - | 241.00, 1937.00 | - | - | - | - |

\*PLWH: People Living With HIV; BMI: body mass index; SBP: systolic blood pressure; DBP: diastolic blood pressure; SD: standard deviation
† PLWH only (n=178)
‡ PLWH who have a detectable Viral Load (n=16)
§ SIM: severe immunosuppression (<200 cells/uL); MIS: moderate immunosuppression (≥200 and <500 cells/uL); NIS: normal immune status (≥500 cells/uL)

The overall study population was predominantly of Black race (97.6%). Males comprised 46.7% of the total study participants, with the proportion varying by group, 32.6% among PLWH, 42.2% among healthy adults, and approximately ranging from 52% to 56% among younger cohorts.

Among the 178 PLWH, 162 participants (91.0%) had an undetectable viral load (<40 copies/mL), whereas 16 (9.0%) exhibited detectable viremia (≥40 copies/mL). Observed values ranged from 55 to 24,000 copies/mL. The CD4 count values ranged from 241 to 1,937 cells/µL with mean and median 843.40 cells/µL (SD = 342.8) and 792.5 cells/µL, respectively. Assessment of the CD4 count showed that none of the participants met criteria for severe immunosuppression (<200 cells/µL). Only 25 participants (14.0%) were categorized as having moderate immunosuppression (200–499 cells/µL), while the majority, 153 individuals (86.0%), demonstrated normal immune status (≥500 cells/µL).

### 3.2. Anti-HEV IgG Seroprevalence

Among the 859 participants who had anti-HEV IgG test results at baseline (before vaccination), 18.04% (155/859) were seropositive and 81.72% (702/859) were seronegative, whereas two participants had equivocal or borderline results for the ELISA test.

#### 3.2.1. Anti-HEV IgG Seroprevalence by Study Site

Site-specific seroprevalence estimates were assessed by sex, age, and HIV status. Since the Mecru Clinical Research Unit site did not enroll children aged 2 – 5 years and 6 – 11 years, site comparisons were limited to adults and adolescents. Significant site-specific variation was observed among healthy adults and adolescents, with higher seroprevalence at Be Part Research (34.5% and 7.3%) and Newtown Clinical Research Centre (23.0% and 14.0%) compared with Mecru Clinical Research Unit (17.2% and 1.5%, p = 0.0499 and 0.0262, respectively) (Table 2, Figure 2).

**Table 2.** Anti-HEV IgG serostatus and seroprevalence by study site, sex, age. [Notes] seropositive rate was compared across sites using Chi-square test [1] or Fisher’s exact test [2]

| Characteristics | Total |  |  | Be Part Research |  |  | Newtown Clinical Research Centre |  |  | Mecru Clinical Research Unit |  |  | P |
| --- | --- | --- | --- | --- | --- | --- | --- | --- | --- | --- | --- | --- | --- |
|  | N | Seropositive, n | Seroprevalence (95% CI) | N | Seropositive, n | Seroprevalence (95% CI) | N | Seropositive, n | Seroprevalence (95% CI) | N | Seropositive, n | Seroprevalence (95% CI) |  |
| Sex |  |  |  |  |  |  |  |  |  |  |  |  |  |
| Male | 401 | 66 | 16.46 (13.15, 20.40) | 146 | 18 | 12.33 (7.94, 18.65) | 165 | 38 | 23.03 (17.26, 30.02) | 90 | 10 | 11.11 (6.15, 19.26) | 0.0119[1] |
| Female | 458 | 89 | 19.43 (16.07, 23.30) | 197 | 48 | 24.37 (18.90, 30.81) | 178 | 31 | 17.42 (12.55, 23.66) | 83 | 10 | 12.05 (6.68, 20.78) | 0.0405[1] |
| HIV Status |  |  |  |  |  |  |  |  |  |  |  |  |  |
| 18-45 years (PLWH) | 178* | 52 | 29.21 (23.03, 36.28) | 66 | 18 | 27.27 (18.00, 39.04) | 66 | 25 | 37.88 (27.15, 49.94) | 46 | 9 | 19.57 (10.65, 33.17) | 0.1009[1] |
| Healthy 18-45 years | 232 | 60 | 25.86 (20.65, 31.86) | 87 | 30 | 34.48 (25.34, 44.94) | 87 | 20 | 22.99 (15.40, 32.86) | 58 | 10 | 17.24 (9.64, 28.91) | 0.0499[1] |
| Age group |  |  |  |  |  |  |  |  |  |  |  |  |  |
| 18-45 years | 410 | 112 | 27.32 (23.23, 31.83) | 153 | 48 | 31.37 (24.55, 39.10) | 153 | 45 | 29.41 (22.77, 37.06) | 104 | 19 | 18.27 (12.02, 26.78) | 0.0525[1] |
| 12-17 years | 174 | 12 | 6.90 (3.99, 11.67) | 55 | 4 | 7.27 (2.86, 17.26) | 50 | 7 | 14.00 (6.95, 26.19) | 69 | 1 | 1.45 (0.26, 7.76) | 0.0262[2] |
| 6-11 years | 175 | 18 | 10.29 (6.61, 15.67) | 85 | 9 | 10.59 (5.67, 18.91) | 90 | 9 | 10.00 (5.35, 17.92) | - | - | - | 0.8981[1] |
| 2-5 years | 100 | 13 | 13.00 (7.76, 20.98) | 50 | 5 | 10.00 (4.35, 21.36) | 50 | 8 | 16.00 (8.34, 28.51) | - | - | - | 0.3724[1] |

\*Two participants - one from Be Part Research and another from Newtown Clinical Research Centre, had equivocal or borderline anti-HEV IgG test results

**Figure 2.**
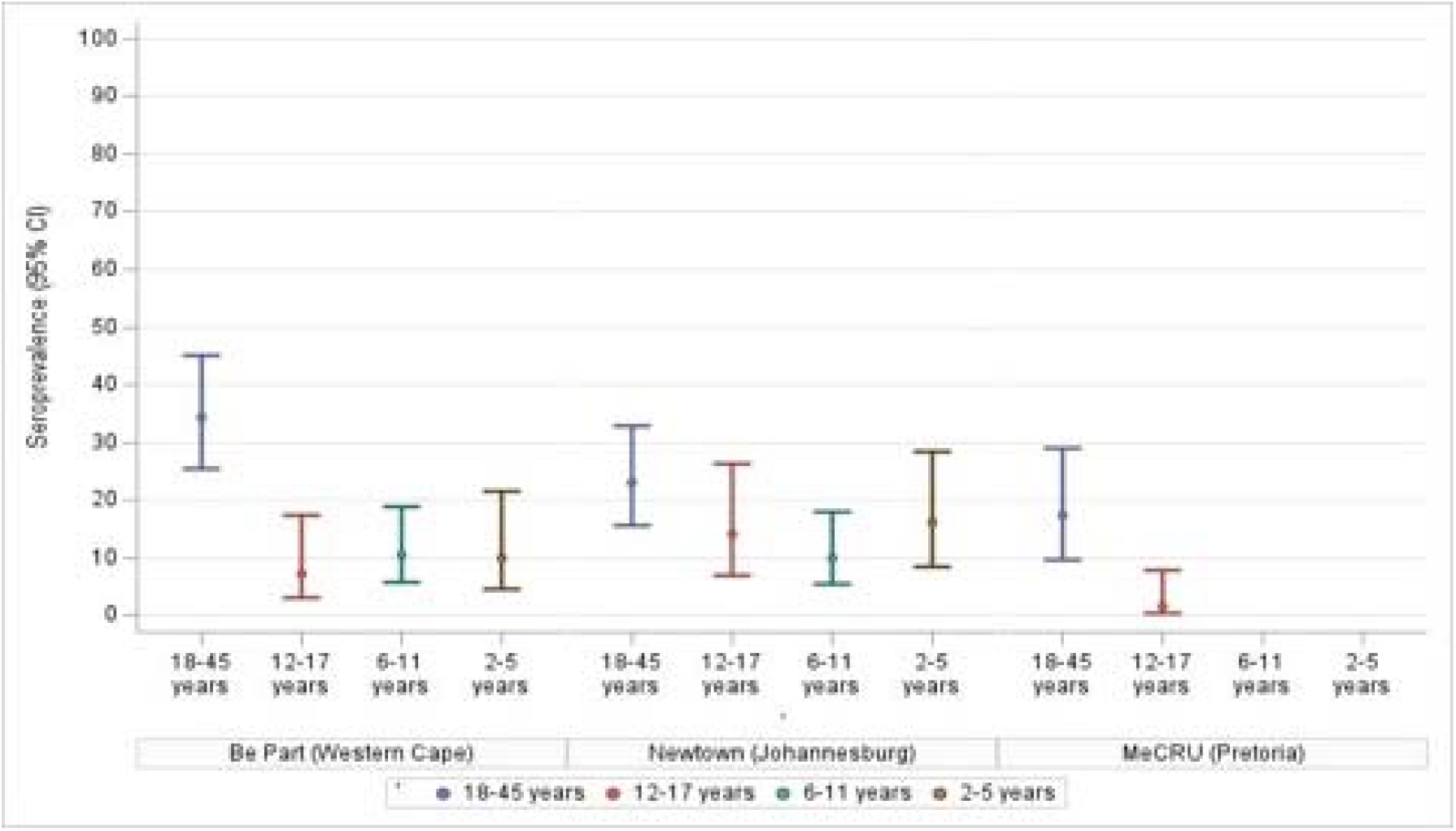
Anti-HEV IgG seroprevalence by site and age group

#### 3.2.2. Anti-HEV IgG Seroprevalence by Age

Overall, the study findings demonstrate an age-related pattern in seroprevalence, highest in adults (25.9% in adults 18–45 years in the general population and 29.2% among PLWH adults), lower in young children (13.0% in those aged 2–5 years and 10.3% in those aged 6–11 years), and lowest among adolescents (6.9% in those aged 12–17 years).

Seroprevalence was markedly lower among children: 6.9% (95% CI: 4.0–11.7) in adolescents (12–17 years), 10.3% (95% CI: 6.6–15.7) in children aged 6–11 years, and 13.0% (95% CI: 7.8–21.0) in those aged 2–5 years (Table 2 and Figure 2). The age-stratified analysis also revealed significant site differences in adolescents aged 12–17 years (*p = 0*.*0262*). Newtown Clinical Research Centre had the highest seroprevalence in adolescents (14.0%), while Mecru Clinical Research Unit had the lowest (1.5%

When all participants stratified by sex, females exhibited a slightly higher seroprevalence (19.4%, 95% CI: 16.1–23.3) compared to males (16.5%, 95% CI: 13.2–20.4). Seroprevalence differed significantly by sex across sites, where among males (n = 401), it ranged from 11.1% at Mecru Clinical Research Unit to 23.0% at Newtown Clinical Research Centre (*p = 0*.*0119*), and among females (n = 458), it ranged from 12.1% at Mecru Clinical Research Unit to 24.4% at Be Part Research (*p = 0*.*0405*) (Table 2).

Seroprevalence was also slightly higher among adult PLWH (29.2%; 95% CI: 23.0–36.3) than among HIV-negative adults (25.9%; 95% CI: 20.7–31.3), but the difference was not statistically significant (Table 2, Figure 3). When stratified by site, adults in the general population (n = 232) had a statistically significant variation across sites (*p = 0*.*0499*), with the highest seroprevalence at Be Part Research (34.5%) and lowest at Mecru Clinical Research Unit (17.2%) (Table 2).

**Figure 3.**
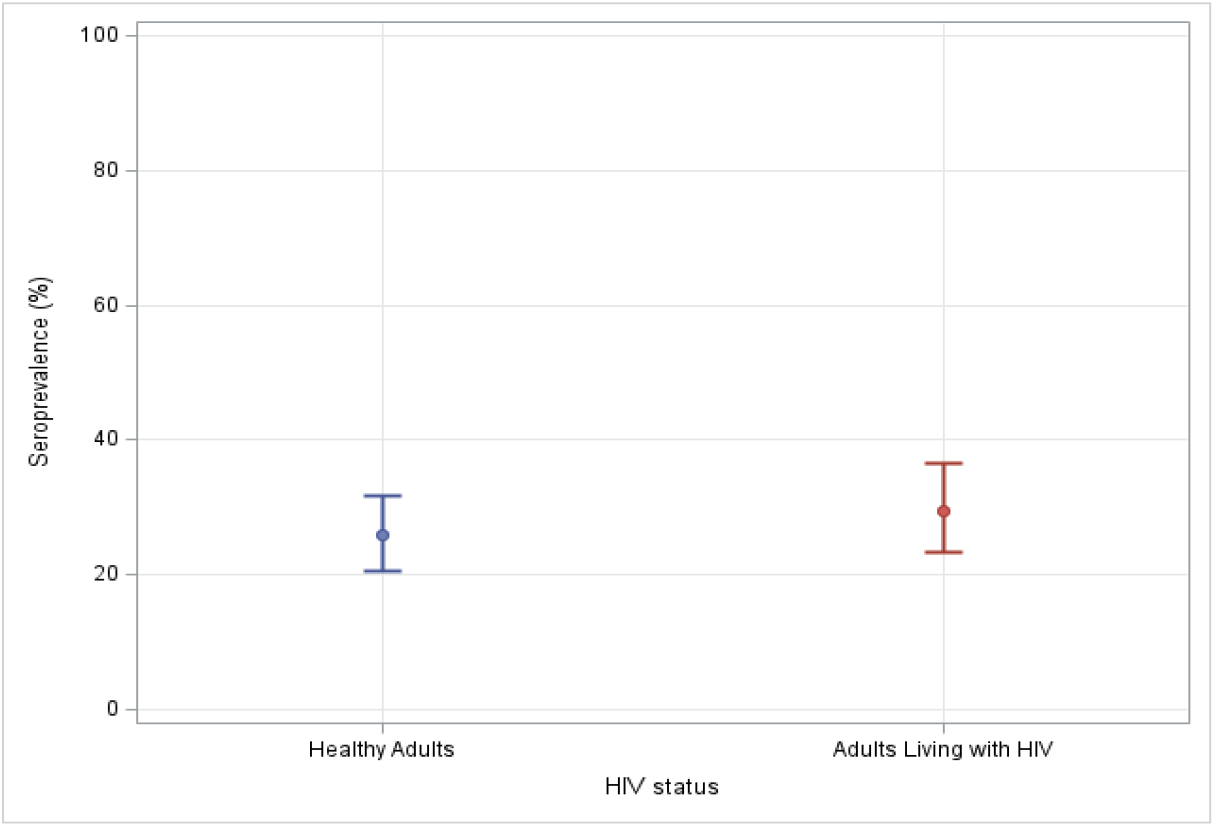
Anti-HEV IgG seroprevalence by HIV status in adult group.

### 3.3. Factors Associated with Anti-HEV IgG Seropositivity

We assessed factors associated with anti-HEV IgG seropositivity among 859 participants, including age, sex, HIV status, virologic markers in PLWH, and site of enrollment (Table 3).

**Table 3.**
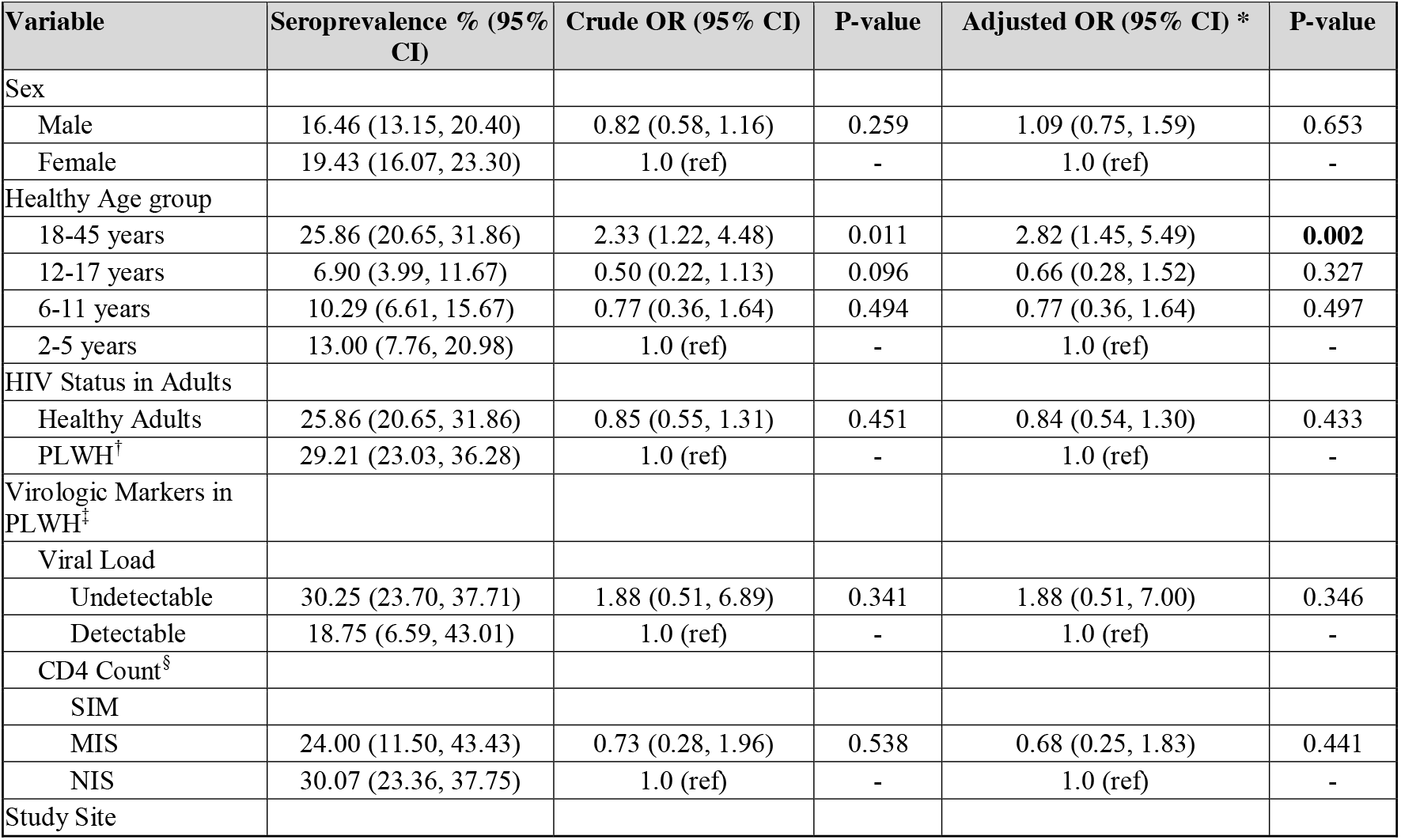
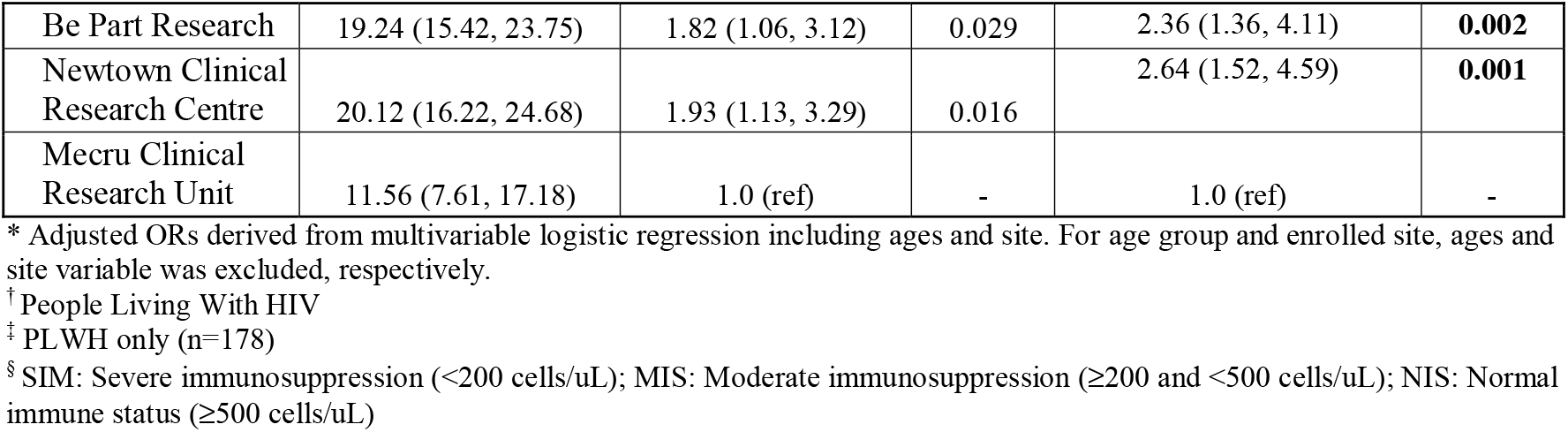
Seroprevalence and crude odds ratios (ORs) with 95% confidence interval (CIs)

| Variable | Seroprevalence % (95% CI) | Crude OR (95% CI) | P-value | Adjusted OR (95% CI) * | P-value |
| --- | --- | --- | --- | --- | --- |
| Sex |  |  |  |  |  |
| Male | 16.46 (13.15, 20.40) | 0.82 (0.58, 1.16) | 0.259 | 1.09 (0.75, 1.59) | 0.653 |
| Female | 19.43 (16.07, 23.30) | 1.0 (ref) | - | 1.0 (ref) | - |
| Healthy Age group |  |  |  |  |  |
| 18-45 years | 25.86 (20.65, 31.86) | 2.33 (1.22, 4.48) | 0.011 | 2.82 (1.45, 5.49) | **0.002** |
| 12-17 years | 6.90 (3.99, 11.67) | 0.50 (0.22, 1.13) | 0.096 | 0.66 (0.28, 1.52) | 0.327 |
| 6-11 years | 10.29 (6.61, 15.67) | 0.77 (0.36, 1.64) | 0.494 | 0.77 (0.36, 1.64) | 0.497 |
| 2-5 years | 13.00 (7.76, 20.98) | 1.0 (ref) | - | 1.0 (ref) | - |
| HIV Status in Adults |  |  |  |  |  |
| Healthy Adults | 25.86 (20.65, 31.86) | 0.85 (0.55, 1.31) | 0.451 | 0.84 (0.54, 1.30) | 0.433 |
| PLWH<sup>†</sup> | 29.21 (23.03, 36.28) | 1.0 (ref) | - | 1.0 (ref) | - |
| Virologic Markers in PLWH<sup>‡</sup> |  |  |  |  |  |
| Viral Load |  |  |  |  |  |
| Undetectable | 30.25 (23.70, 37.71) | 1.88 (0.51, 6.89) | 0.341 | 1.88 (0.51, 7.00) | 0.346 |
| Detectable | 18.75 (6.59, 43.01) | 1.0 (ref) | - | 1.0 (ref) | - |
| CD4 Count<sup>§</sup> |  |  |  |  |  |
| SIM |  |  |  |  |  |
| MIS | 24.00 (11.50, 43.43) | 0.73 (0.28, 1.96) | 0.538 | 0.68 (0.25, 1.83) | 0.441 |
| NIS | 30.07 (23.36, 37.75) | 1.0 (ref) | - | 1.0 (ref) | - |
| Study Site |  |  |  |  |  |
| Be Part Research | 19.24 (15.42, 23.75) | 1.82 (1.06, 3.12) | 0.029 | 2.36 (1.36, 4.11) | **0.002** |
| Newtown Clinical Research Centre | 20.12 (16.22, 24.68) | 1.93 (1.13, 3.29) | 0.016 | 2.64 (1.52, 4.59) | **0.001** |
| Mecru Clinical Research Unit | 11.56 (7.61, 17.18) | 1.0 (ref) | - | 1.0 (ref) | - |

\* Adjusted ORs derived from multivariable logistic regression including ages and site. For age group and enrolled site, ages and site variable was excluded, respectively.
† People Living With HIV
‡ PLWH only (n=178)
§ SIM: Severe immunosuppression (<200 cells/uL); MIS: Moderate immunosuppression (≥200 and <500 cells/uL); NIS: Normal immune status (≥500 cells/uL)

Seroprevalence was slightly higher among females (19.4%) than males (16.5%), but there was no statistically significant association in either crude (OR 0.82, 95% CI: 0.58–1.16, *p = 0*.*259*) or adjusted analyses (aOR 1.09, 95% CI: 0.75–1.59, *p = 0*.*653*).

Compared with the reference group of children aged 2–5 years, adults aged 18–45 years had significantly higher odds of seropositivity (crude OR 2.33, 95% CI: 1.22–4.48, *p = 0*.*011*; aOR 2.82, 95% CI: 1.45– 5.49, *p = 0*.*002*). Adolescents (12–17 years) and children aged 6–11 years did not show significant differences compared with the 2–5 years reference group.

Seroprevalence was slightly higher among PLWH adults (29.2%) compared with healthy adults (25.9%), but the difference was not statistically significant (aOR 0.84, 95% CI: 0.54–1.30, p = 0.433). Virologic markers among PLWH: Adults with undetectable viral load had a higher seroprevalence (30.3%) than those with detectable viral load (18.8%), but the association was not statistically significant (aOR 1.88, 95% CI: 0.51–7.00, *p = 0*.*346*). CD4 count categories (SIM, MIS, NIS) were also not significantly associated with seropositivity.

Compared with Mecru Clinical Research Unit (reference, 11.6%), participants from Be Part Research (19.2%) and Newtown Clinical Research had significantly higher odds of seropositivity in both crude (Be Part Research OR 1.82, *p = 0*.*029*; Newtown Clinical Research Centre OR 1.93, *p = 0*.*016*) and adjusted analyses (Be Part Research aOR 2.36, *p = 0*.*002*; Newtown Clinical Research Centre aOR 2.64, *p = 0*.*001*).

### 3.3. Anti-HEV IgG Antibody Titer

Adult PLWH had a mean anti-HEV IgG titer of 0.68 IU/mL (SD 0.96), comparable to HIV-negative adults of the same age (0.88 IU/mL, SD 1.54). Adolescents aged 12–17 years had a mean titer (0.67 IU/mL, SD 0.94), while young children demonstrated higher titers, with mean levels of 2.02 IU/mL (SD 5.68) in those aged 6–11 years and 5.06 IU/mL (SD 6.53) in the 2–5-year group (Figure 4). These findings indicate significantly higher anti-HEV IgG titers in younger children compared to adolescents and adults, suggesting age-related differences in antibody responses or more recent infection. The observed age-specific difference in anti-HEV IgG antibody titers was statistically significant (*p = 0*.*0371*).

**Figure 4.**
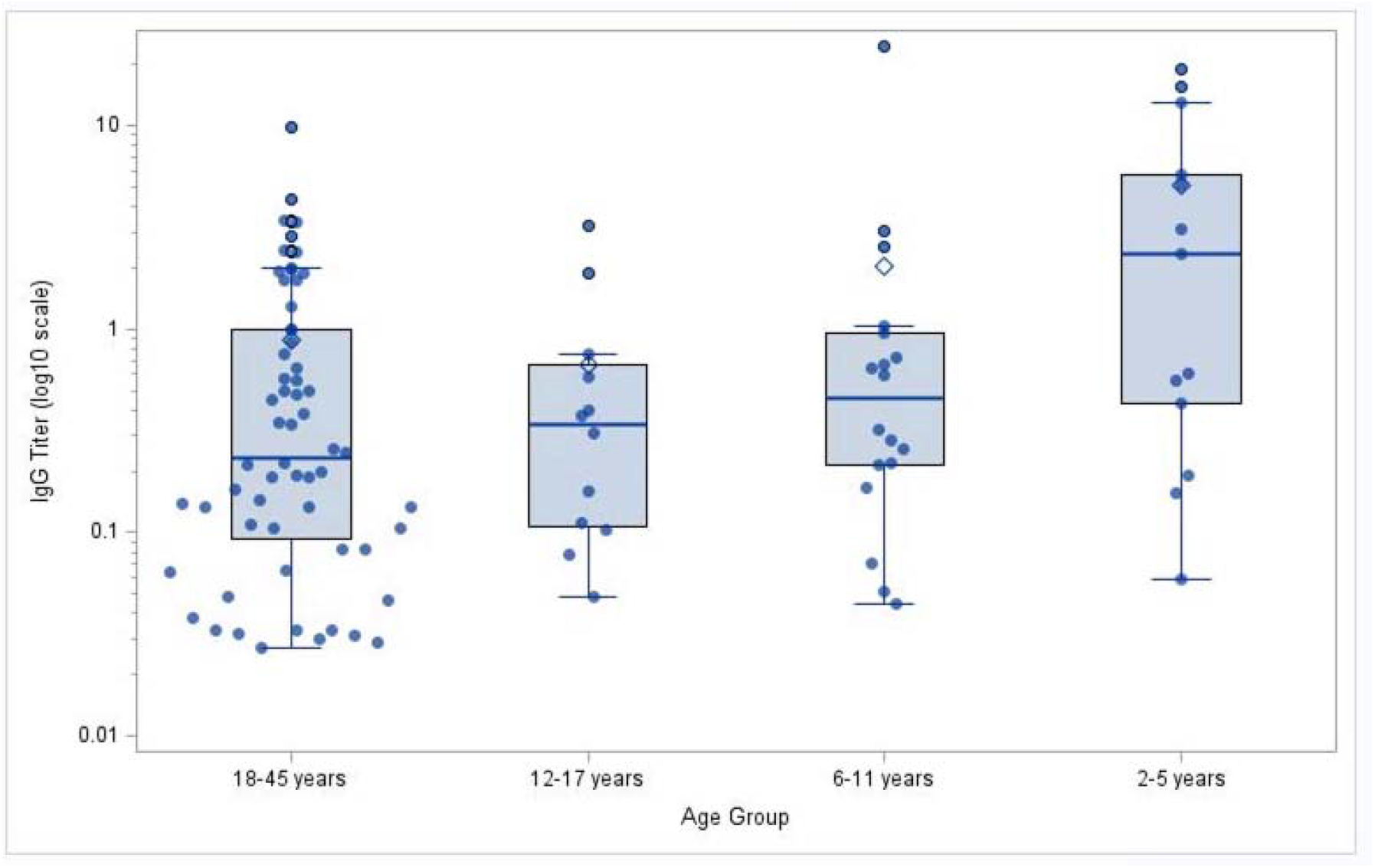
Anti-HEV IgG titer at baseline by age group in healthy seropositive participants.

## 4. Discussion

In this study, we documented an overall anti-HEV IgG seroprevalence of 18.0% (95% □CI:□15.6–20.8) among participants comprising children, adolescents, HIV-negative adults, and adult PLWH enrolled at three clinical sites in South Africa. Our findings reveal heterogeneity by age, HIV status, sex, and geographic location, underscoring a considerable variation in HEV epidemiology within South Africa. This moderate seroprevalence aligns with several African studies (26), but is lower than the highest reported rates on the continent including reports from South Africa (19, 20, 23), suggesting different factors play key roles in shaping HEV exposure patterns.

The 18.0% (95% □CI□15.6–20.8) overall estimated seroprevalence in our study is lower than the 27.9% (95% CI 25.3–30.5) reported from the Western Cape Province by Madden *et al*. (2016), despite both studies employed the same antibody detection assay, Wantai HEV IgG ELISA. Even when focusing specifically on our site located in Mbekweni, Western Cape Province, the seroprevalence estimate of 19.2% remains below the previously reported figure(19). Differences in cohort age structure, geographic context, and characteristics likely contribute to this discrepancy. The inclusion of a large paediatric cohort, who had the lowest seroprevalence, contrasts with the more adult-weighted population in the Western Cape study, where seropositivity rose sharply after adolescence (19).

We observed substantial geographic variation in anti-HEV IgG seroprevalence, with adult and adolescent participants from Johannesburg (29.4% and 14.0%) and Mbekweni, Western Cape (31.4% and 7.3%) showing higher seroprevalence compared to Garankuwa, Pretoria (18.3% and 1.5%, respectively). Similar variation has historically been documented in South Africa – early work demonstrated sporadic HEV circulation with higher seroprevalence in rural communities (21), while more recent reports have shown variation across provinces (20, 27). A study among patients with acute hepatitis in Western Cape reported an anti-HEV IgG prevalence of approximately 29.5% (23), and the other study in the Western Cape on a comprehensive regional analysis confirmed an age-adjusted seroprevalence of 21.9% and identified pork consumption as a key exposure linked to zoonotic HEV genotype 3 transmission (19).

Environmental surveillance data from Salemane *et al*. (2024) identified HEV RNA in approximately 21.8% of wastewater and surface water samples across South African provinces, with particularly high detection in regions such as the Western Cape (38.1%) (28). Historical seroepidemiologic data also showed high anti-HEV IgG seroprevalence (28% to 43%) in the Western Cape (23, 29). In contrast, a seroprevalence study conducted near Pretoria found a low HEV exposure rate of 3.1% in HIV negative pregnant women, suggesting limited local HEV transmission (27). These patterns may suggest that HEV exposure rate could be greater in Western Cape and Johannesburg compared to Pretoria.

In contrast, a 2024 study conducted in the Free State, South Africa found an exceptionally high IgG seroprevalence of 60.9%, suggesting a distinct epidemiological profile in that province (20). Earlier serological study in specific South African populations demonstrated wide variability, with seroprevalence estimates influenced by local environmental and behavioural exposures (22). Collectively, these findings highlight that HEV exposure in South Africa is not uniform but shaped by region-specific factors which mirrors global patterns of substantial heterogeneity driven by differences in sanitation, water quality, dietary practices, and animal reservoir distribution. These findings underscore the importance of multi-site sampling, regional risk assessment, and tailored public health strategies for accurately characterizing and addressing HEV epidemiology.

Overall, these findings highlight the value of multi-site sampling for estimating HEV epidemiology in South Africa, as exposure risk can vary by location – a pattern consistent with region-specific heterogeneity observed across Africa and globally. For example, in Uganda’s Rakai District, a community-based serosurvey found anti-HEV IgG seroprevalence of 47% among adults, signifying a very high burden of past HEV exposure in that rural setting (30). A broader systematic review of HEV in Africa similarly documented variation in seroprevalence in different countries ranging from as low as 0% in some rural equatorial African villages residents in Gabon (26, 31) to extremely high rates of up to 84.3% in pregnant women in Egypt (26, 32). Another meta-analysis focused on pregnant women across Africa estimated a pooled seroprevalence of 29.1% (95% CI 14.6–43.6%), with highest rates in North and East Africa and substantial heterogeneity by assay and geography (18). Zoonotic contributions are also likely important: a systematic review of HEV in African animals reported a pooled IgG seroprevalence of 23.4%, with pigs showing particularly high rates (35.1%), underscoring the potential for cross-species transmission (33).

In this multi-site study, we also observed a clear age-dependent pattern in anti-HEV IgG seroprevalence, where adults had the highest seroprevalence, particularly PLWH (29.2%), followed by HIV-negative adults (25.9%), while children and adolescents had substantially lower seroprevalence rates. Our observations are partially consistent with those reported in the Western Cape Province of South Africa study, who also reported an increase in seroprevalence with age, from approximately 10% in children (<19 years) to nearly 30% in adults aged 30–39 years and plateaued thereafter (19). Another study from Western Cape also found a seroprevalence increase with age: 5% (≤21 years), 26% (22 – 45 years), and 46% (>46 years) (p< 0.001) (34). In Malawi, Taha *et al*. (2015) reported a 16.5% adult seroprevalence, indicating that exposure accumulates primarily in adulthood rather than early childhood (35). The age-related seroprevalence pattern is even more pronounced in a Zambian study by Jacobs *et al*. who reported that seroprevalence rose sharply from 8% in children aged 1–4 years to 36% by ages 10–14, eventually reaching 42% in adults, with rates as high as 71% among PLWH (10). Compared with Zambia, our study shows delayed acquisition of the HEV, with relatively lower seroprevalence in adolescents and a more pronounced rise only in adulthood.

However, recent longitudinal serological data from a population-representative cohort in Bangladesh indicate that interpretation of such age patterns may be accounted for by antibody waning. Over approximately 9 months of follow-up, about 15% of participants who were initially anti-HEV IgG positive lost detectable antibodies annually, and seroreversion was significantly more common in children than in adults. Most seropositive children below 10 years became seronegative within months, compared with much longer persistence in older age groups (36). This rapid waning of detectable antibodies in childhood could contribute to underestimation of early exposure in cross-sectional studies and produce an apparent trough in seroprevalence during adolescence, even when early life infections have occurred.

Therefore, the low adolescent seroprevalence we observe may be partly due to antibody kinetics rather than solely lower exposure, suggesting that infection incidence in younger age groups might be higher than cross-sectional data imply. Accounting for seroreversion is important because ignoring waning can substantially underestimate the true infection risk derived from seroprevalence alone.

The relatively low seroprevalence observed among adolescent participants (12–17 years) in our study may reflect a transitional period in which antibody from early infection wanes and exposure risk temporarily declines, possibly due to the fact that adolescents have a lower risk compared with adults who may have occupational exposures (e.g., occupational animal contact or consumption of higher-risk foods) or effects arising from improvements in hygiene and food safety compared to the young children. These interpretations, however, should be made cautiously, as cross-sectional data cannot disentangle true differences in exposure from antibody waning (36) or generational effects (37).

In our study, the anti-HEV IgG seroprevalence was slightly higher in females (19.4%, 95% CI: 16.1– 23.3) than in males (16.5%, 95% CI: 13.2–20.4), but this difference was not statistically significance (OR = 0.82, 95% CI: 0.75–1.59; p = 0.653) suggesting that sex is not a strong independent predictor of past HEV exposure in South Africa. This aligns with the previous studies done in South Africa: for instance, the recent study in the Free State province of South Africa, which reported more males (63.3%) than females (59.5%) were seropositive, but found no statistically significant difference in seropositivity by sex (p = 0.81) (20). Similarly, the study from Western Cape of South Africa, reported no significant difference by gender (23). However, studies in other settings and populations such as a study among PLWH in Central African Republic and a cohort study of PLWH in Nepal found significantly higher seroprevalence among women, suggesting that gender related differences in exposure risk may be more prominent in certain populations or socio-cultural settings (38).

Among our adult participants, PLWH exhibited a slightly higher anti-HEV IgG seroprevalence (29.2%, 95% CI: 23.0–36.3) than HIV-negative adults (25.9%, 95% CI: 20.7–31.9), but the difference was not statistically significant (OR = 0.85, 95% CI: 0.55–1.31; *p = 0*.*451*). This is consistent with prior epidemiological evidence including a large meta-analysis of immuno-compromised populations including HIV-positive patients which found no significant difference in anti-HEV IgG seroprevalence between HIV-infected individuals and other immunosuppressed groups such as transplant recipients (39). The cross-sectional study in the Rakai District, Uganda, involving 500 HIV-positive adults and matched HIV-negative individuals, observed a very high anti-HEV IgG seroprevalence (47%) but found no association between HIV status and anti-HEV IgG seropositivity (prevalence ratio 0.97, 95% CI: 0.85–1.11) (30).

However, in Zambia, a study found a strong association between HIV infection and HEV seroprevalence, where HIV was significantly associated with anti-HEV IgG positivity in adults, although the study population included individuals with very low CD4 counts (10). The inconsistency of the association across settings and populations again suggests that HIV infection per se may not dramatically alter the likelihood of HEV exposure.

Quantitative IgG measurements in our study provided important additional insight into the timing and intensity of HEV exposure across age groups and sites, where young children (2–5 years) had the statistically significant highest mean titres (5.06 IU/mL), whereas adults exhibited lower titres despite higher seroprevalence. This may suggest that younger children’s infections were more recent and that antibody levels may decline over time following exposure. This pattern is consistent with general seroepidemiologic understanding that antibody titers can reflect the recency and magnitude of exposure.

## Conclusions

In conclusion, our findings reveal a moderate overall anti-HEV IgG seroprevalence (approximately 18%), across these three geographic settings in South Africa. We observed an age-related seroprevalence pattern, characterized by the highest seroprevalence in adults, the lowest in adolescents, and comparatively high antibody titers in young children (2–5 years). These findings suggest recent exposure among those 2-5 years with declining seroprevalence and titers in later childhood and adolescence, followed by rising seroprevalence in adults through cumulative new exposures. The lower prevalence observed among adolescents may represent a transitional phase with reduced exposure and/or waning antibody levels following early childhood infection, although this warrants further investigation.

Seroprevalence also varied by site, suggesting that local environmental, sanitary, sociocultural or dietary factors may play an important role in shaping exposure risk. HIV status, viral load, and CD4 count had limited impact on seropositivity among adults. These findings provide valuable insights into HEV epidemiology in South Africa and can inform vaccination and public health strategies, particularly in regions with heterogeneous exposure patterns.

## Data Availability

De–identified data underlying the results reported in this article will be made upon submission and approval of a formal data access request

## 5. Strengths and Limitations

Strengths of our study include a relatively large sample size drawn from multiple sites in South Africa with broad age group inclusion and including PLWH. In addition, this study includes quantitative assessment of anti-HEV IgG titers with all testing performed at a centralized lab using an international reference standard. Limitations include that the participants were not randomly selected from the communities but were volunteers in a clinical trial which may introduce selection bias. In addition, the cross-sectional nature of sampling cannot distinguish recent from past infections and no HEV genotyping or extended risk factor for exposure could be assessed beyond geographic residence, age, sex and HIV status.

## Acknowledgements

The authors thank and recognize the children, their parents, and adult participants who made this study possible, and the Be Part Research (Mbekweni, Paarl in Western Cape), Newtown Clinical Research Centre (in Johannesburg), and Mecru Clinical Research Unit) sites study staff for their dedication in conducting the study. We also thank Haekyoung Park, the project administrator at International Vaccine Institute, for providing excellent administrative assistance. We acknowledge the Cytespace Laboratories staff (in Pretoria) for their collaboration in setting up the assays and performing the laboratory tests. In addition, the International Vaccine Institute acknowledges Xiamen Innovax Biotech Co., Ltd. China and Beijing Wantai Biological Pharmacy Enterprise, Beijing, China for donating the Wantai HEV IgG ELISA kits.

## 8. Author contributions

JAL, TS, NT, EH, EM, MN, RB, ND, and SA developed the study design. TS, NT, EH, EM, MN, RB, ND, SA, SV, SP, and JAL supervised the sample and data collection and analysis. JP, LY, JL, and DK managed the database and performed statistical analysis. NT wrote original draft, and JAL, TS, JY, SA, EH, EM, MN, RB, ND, HP, and JS reviewed and edited the manuscript. All authors have read and approved the final version of the manuscript.

